# COVID-19 Pandemic and Academic Speculation of Medical Students of Bangladesh: A Cross-sectional, Comparative Study

**DOI:** 10.1101/2021.05.11.21257042

**Authors:** Fatema Johora, Asma Akter Abbasy, Fatiha Tasmin Jeenia, Mithun Chandro Bhowmik, Priyanka Moitra, Sabiha Mahboob, Jannatul Ferdous

## Abstract

**Background:** COVID-19 pandemic has caused unprecedented disruptions worldwide including education system. While the necessary focus has been on patient care andwellbeing of healthcare professionals, the impacts on medical students need to be discussed.

**Methods:** This cross-sectional comparative study was conducted to evaluate the academic speculation of medical students studying in government and non-government institute during COVID-19 pandemic. A structured questionnaire survey linked in the google form was used as study instrument and was distributed among study population through email, messenger, whatsapp and other social media. Total 1020 students were participated in the study

**Results:** In this research, 441 (43.24%) and 579 (56.77%) students were from government and non-government medical colleges respectively. Opinion of both group was almost similar regarding disruption of medical education, loss of clinical skills and competency, future career plan, and stress and anxiety but significant differences were observed between both group regarding issue of financial burden, meaningful learning opportunities, fear of getting infected and maintenance of social distancing in hostel.

**Conclusion:** The study revealed similar kind of viewpoint about disruption of education, loss of clinical skill and competency, changing aspects of future career plan and increased level of stress and anxiety among medical students of government and non-government institutes of Bangladesh but different speculations were found regarding issue of financial burden, meaningful learning opportunities, fear of getting infected and possibility of maintenance of social distancing in hostel.

## Introduction

COVID-19 emerged in Wuhan at the end of December 2019 and the disease spread quickly worldwide. To limit the spread of COVID-19 and avoid overburdening the healthcare system, several countries have implemented various forms of lockdown including social distancing, self-quarantine or isolation, closing public facilities, including institutions, museums, places for large gatherings, which included universities and schools.^[1]^ Over 91% of students all around the world are affected by the closing of education institutions in their countries.^[2,3]^ In total, there are more than 900 million learners have been affected.^[4]^ In this unprecedented time, almost all sectors are uncertain on how to act including the medical education field. It’s high time for preparing future physicians as this global pandemic hits and yet practical, ethical, and logistical challenges exist as medical students may potentially spread or acquire the virus during training, thereby endangering both themselves, their family, or other patients.^[5]^ Since the institutions that provide medical education are deeply affected by the pandemic, medical educators were in need to find new ways to keep the students engaged in their education. Several countries took different measures in continuing medical education.Some universities have shifted face-to-face classes to online education to provide various courses and programs, and some have replaced the live clinical exposure to the virtual one. ^[6,7]^

Like many other countries, the government of Bangladesh decided to close the educational institutions as part of preventive measures against the spread of COVID-19 pandemic.In Bangladesh, total 112 medical colleges (36 public medical colleges, 70 private medical colleges and 06 medical colleges run by Bangladesh Armed Forces) are assigned to provide the formal Medical education with the aim of making graduate competent under the guidance of BMDC. ^[8]^ In view of the prolonged closure of institutions and the uncertainty of re-opening, Bangladesh Government switched onto online education from conventional face-to-face education.However, virtual teaching being a new experience in almost all medical colleges; sudden shifting of teaching methodology has imposed challenges to both the faculty and students in adopting the new virtual class^. [9,10]^

Hence, the present study was carried out with the attempt to find out and compare theacademic speculation of medical students of Bangladesh studying in government and non-government institute during COVID-19 pandemic.

## Material &Methods

The objective of the study was to find out and compare academic speculation of medical students of Bangladesh studying in government and non-government institute during COVID-19 pandemic.

### Study Design and Population

A cross-sectional comparative study was designed to meet the study objective. The study population comprised of 3rd, 4th and final year students of seven medical colleges of Bangladesh including government (Colonel Malek Medical College, Manikganj and Rangpur Medical College) and non-government medical colleges (Army Medical College Bogura, Army Medical College Chattogram, BGC Trust Medical College, Brahmanbaria Medical College and Chattogram International Medical College) in October 2020.

### Study Instrument

A structured questionnaire consisted of 08 Likert scale (five-point) questions was used for data collection and questionnaire was validated before survey.

### Procedure

Permission was taken from college authorities and informed consent was taken from the participants of the Structured Questionnaire Survey.Researchers explained the nature and purpose of the survey to the students during a virtual class. This self-administered questionnairewas linked in google form and was distributed amongstudy population through email, messenger, whatsapp and other social mediawho gave consent. To assure the quality, students filled and submitted the questionnaire quickly during theend of class. Later, this web-based questionnaire was sent to students who were absent in the class through email.A reminder mail or message wasgiven on 7^th^ day and 15^th^ day of the primary one.The responses generated by the students were received through google drive,and it did not accept double response from same participant.

### Statistical analysis

Data was compiled, presented and analyzed using SPSS version 22, and was expressed as percentage and mean values. Unpaired t-test was done to determine the significance of difference between the mean values. Statistical analysis was performed at a 95% confidence interval and significance was determined at p< 0.05.

## Results

One thousand and twenty respondents were covered during the study period, of which 330 (32.35%) were males and 690 (67.64%) were females. Students from government and non-government medical colleges were 441 (43.24%) and 579 (56.77%) respectively (Table 1).

**Table 1:** Demographic Information.

| Demographic Information | Particulars | Frequency | Percentages |
| --- | --- | --- | --- |
| Gender | Male | 330 | 32.35% |
|  | Female | 690 | 67.64% |
| Types of Institute | Govt. | 441 | 43.24% |
|  | Non-govt. | 579 | 56.77% |

**Table 2** showed that among the students of government medical colleges, majority (56.01%) of the students strongly agreed that their medical education has been significantly disrupted by the pandemic, majority (50.5%) agreed that pandemic is going to limit their clinical skills and competency and it will influence the career plan of many (48.3%) of them. Most (56.46%) of the students strongly agreed that their academic career is going to be lengthen by this pandemic which will increases the financial burden of their family and many (41.72%) of them disagreed on the issue of finding a meaningful learning from this pandemic. Rather many of them (44.22%) strongly agreed that their stress and anxiety level has been increased. Though most (53.06%) of the students disagreed about the feasibility of maintaining social distancing in classroom, ward and hostel, still many of them (39.46%) agreed strongly to accept the risk of getting infected with COVID 19 while returning to clinical ward placement.

**Table 2:** Responses of students studying in government insitute to the questionnaire.

| Question | Response (N=441) |  |  |  |  |  |  |  |  |  |
| --- | --- | --- | --- | --- | --- | --- | --- | --- | --- | --- |
|  | Strongly agree |  | Agree |  | Neither agree or disagree |  | Disagree |  | Strongly disagree |  |
|  | N | % | N | % | N | % | N | % | N | % |
| My medical education has been significantly disrupted by the pandemic | 247 | 56.01 | 187 | 42.4 | 4 | 0.91 | 3 | 0.68 | 0 | 0 |
| The pandemic is going to limit my clinical skills and competency | 192 | 43.54 | 223 | 50.57 | 18 | 4.08 | 8 | 1.81 | 0 | 0 |
| COVID 19 will definitely influence my future career plan | 174 | 39.46 | 213 | 48.3 | 30 | 6.8 | 20 | 4.54 | 4 | 0.91 |
| As this pandemic is going to lengthen my academic career, it would be a financial burden for my family | 249 | 56.46 | 158 | 35.83 | 22 | 4.99 | 10 | 2.27 | 2 | 0.45 |
| I have been able to find meaningful learning opportunities in the pandemic | 27 | 6.12 | 83 | 18.82 | 96 | 21.77 | 184 | 41.72 | 51 | 11.56 |
| My stress and anxiety level has been increased in this pandemic | 195 | 44.22 | 183 | 41.5 | 23 | 5.22 | 37 | 8.39 | 3 | 0.68 |
| It is possible to maintain social distancing in classroom, ward and hostel. | 12 | 2.72 | 39 | 8.84 | 10 | 2.27 | 234 | 53.06 | 16 | 33.11 |
| I accept the risk that I may be infected with COVID 19, if I return to clinical ward placement | 174 | 39.46 | 169 | 38.32 | 24 | 5.44 | 46 | 10.43 | 28 | 6.35 |

Table 3 showed that among the students of non-government medical colleges, majority (65.63%) of the students strongly agreed that their medical education has been significantly disrupted by the pandemic and it is going to limit the clinical skills and competency of many of them (47.84%). About (44.73%) of them agreed that it will influence their career plan. Mostof the students (75.13%) strongly agreed that their academic career is going to be lengthen by this pandemic which will increases the financial burden of their family and many (35.75%) of them disagreed on the issue of finding a meaningful learning from this pandemic. Rather almost half of the respondents (50.6%) strongly agreed that their stress and anxiety level has been increased. Though many respondents (43.01%) disagreed about the feasibility of maintaining social distancing in classroom, ward and hostel, still they (43.01%) agreed to accept the risk of getting infected with COVID 19 while returning to clinical ward placement.

**Table 3:** Responses of students studying in non-government insitute to the questionnaire.

| Question | Response(N= 579) |  |  |  |  |  |  |  |  |  |
| --- | --- | --- | --- | --- | --- | --- | --- | --- | --- | --- |
|  | Strongly agree |  | Agree |  | Neither agree or disagree |  | Disagree |  | Strongly disagree |  |
|  | N | % | N | % | N | % | N | % | N | % |
| My medical education has been significantly disrupted by the pandemic | 380 | 65.63 | 170 | 29.36 | 11 | 1.9 | 17 | 2.94 | 1 | 0.17 |
| The pandemic is going to limit my clinical skills and competency | 277 | 47.84 | 267 | 46.11 | 22 | 3.8 | 12 | 2.07 | 1 | 0.17 |
| COVID 19 will definitely influence my future career plan | 243 | 41.97 | 259 | 44.73 | 43 | 7.43 | 29 | 5.01 | 5 | 0.86 |
| As this pandemic is going to lengthen my academic career, it would be a financial burden for my family | 435 | 75.13 | 117 | 20.21 | 14 | 2.42 | 9 | 1.55 | 4 | 0.69 |
| I have been able to find meaningful learning opportunities in the pandemic | 37 | 6.39 | 141 | 24.35 | 132 | 22.8 | 207 | 35.72 | 62 | 10.71 |
| My stress and anxiety level has been increased in this pandemic | 293 | 50.6 | 211 | 36.44 | 38 | 6.56 | 32 | 5.53 | 5 | 0.86 |
| It is possible to maintain social distancing in classroom, ward and hostel. | 50 | 8.64 | 66 | 11.4 | 39 | 6.74 | 249 | 43.01 | 175 | 30.02 |
| I accept the risk that I may be infected with COVID 19, if I return to clinical ward placement | 160 | 27.63 | 249 | 43.01 | 43 | 7.43 | 64 | 11.05 | 63 | 10.88 |

**Table 4** showed that significance differences were present between government and non-government students regarding financial impact, meaningful learning opportunities, possibility of maintenance of social distancing and probability of getting infected.

**Table 4:** Comparison between the responses of students studying in government and non-government medical colleges.

| Question | Response |  |  |
| --- | --- | --- | --- |
| | Government<br>Mean ± SD<br>(n=441) | Non-government<br>Mean ± SD<br>(n=579) | P value |
| My medical education has been significantly disrupted by the pandemic | 4.54± .56 | 4.57± 0.69 | 0.46 |
| The pandemic is going to limit my clinical skills and competency | 4.36±0.65 | 4.39± 0.68 | 0.48 |
| COVID 19 will definitely influence my future career plan | 4.21± 0.83 | 4.22± 0.85 | 0.86 |
| As this pandemic is going to lengthen my academic career, it would be a financial burden for my family | 4.46± 0.73 | 4.68± 0.67 | < 0.0001 |
| I have been able to find meaningful learning opportunities in the pandemic | 2.66± 1.10 | 2.80±1.12 | 0.0465 |
| My stress and anxiety level has been increased in this pandemic | 4.20±0.92 | 4.30± 0.88 | 0.078 |
| It is possible to maintain social distancing in classroom, ward and hostel. | 1.95±0.98 | 2.25±1.24 | < 0.0001 |
| I accept the risk that I may be infected with COVID 19, if I return to clinical ward placement | 3.94± 1.20 | 3.65±1.29 | 0.0002 |

Unaired t-test was done; $P \leq 0.05$ = Statistically significant
1= Strongly disagree, 2= Disagree, 3= Neither agree or disagree, 4= Agree, 5= Strogly agree

## Discussion

COVID-19 pandemic has been challenging healthcare and medical education globally. Current study was conducted in this context to assess and comapre academic speculations of medical students of Bangladesh studying in government and non-government institutions during COVID-19 pandemic.

Medical education is a dynamic as well as complex process, commences at undergraduate level and continues until a physician retires from active practice. The years spent in medical school are formative for future physicians as it is expected that medical schools should equip the students with knowledge, attitude and skills required for the clinical practice throughout theirlife.^[11]^In the last few months, COVID-19 pandemic has been deleteriously affectingevery aspects of human life including education and healthcare system.^[12]^Closing of health institutions, distance learning tactics, compliance to virtual learning, loss of clinical experience and classroom learning experience, no proper examination, disruption of professional development all leads to a disruption in the education of future physicians.^[13,14,15]^Although medical schools in Bangladesh adapted online teaching-learning from the very beginning of the pandemic,students of both govt. and non-govt. medical collegesagreed about disruptionof ongoing learning process.^[9, 16]^ They also stated that this learning disruption will limit their clinical skills and competency as many of them have missed exposure to entire specialties; that they may now never have the opportunity to experience.^[17]^ This will have a profound impact, not only on knowledge in these areas but also on progression through medical college and career choice.^[18]^The participants of current research also agreed that COVID-19 pandemic will definitely influence their future career plan and similar kind of viewpoint was observed in a study conducted in US.^[19]^

COVID-19 pandemic emerged as the most devastating and challenging crisis for public health in the contemporary world. Apart from the high mortality rate, nations across the globe have also been suffering from a spike of the excruciating psychological outcomes, i.e., anxiety and depression among people of all ages.^[19,20,21]^Sudden changes in curricular delivery, struggling with virtual learning, fear of lossing clinical skill, loss of peer interaction, social isolation, uncertainity, ambiguity around future prospects and subsequent stree all are negatively affecting mental well-being of medical students and similar kind of anxiety and stress were perceived by medical students of Bangladesh.^[20, 21, 22, 23]^

Although COVID-19 pandemic has been considered as a source of disruption, it is likely that it will also be viewed as a catalyst for the transformation of medical education as the pandemic has provided an opportunity for learners to realize the dynamic nature of medical knowledge and medical students not onlyabout continued learning through alternative ways but in many circumstances, also contributing efforts to reduce the impact of the pandemic and accelerating their attainment of competencies that 21^st^ century needs.^[5,15,16,24]^In this study, most of the respondentsdisagreed about opportunities regarding learning new things and statement provided by students of government and non-government institute was found statistically significant (2.66± 1.10vs2.80±1.12, p 0.0465). Less familiarity with virtual learning along with cost of smart phone, laptop, internet might be act as barriers in learning process.^[15,16,24]^And this issue was more observed among thestudents of goverment medical college, probably due to less financial solvency.

Medical education is expensive worldwide. Digital device or infrastructure required for online learning putting extra cost on medical students belonging to low socioeconomic status.^[15,16]^ Prolongation of course length due to postponing of exams for an indefinite period can have significant financial implications and it was expressed by participants of this study. Students from non-govt. medical colleges were more concerned about financial implications of COVID-19 pandemic as in Bangladesh (4.46± 0.73 vs 4.68± 0.67, p < 0.0001), higher education in non-government institute is costly.^[25,26]^Most of the studentsfeared of getting infected with coronavirus when they will return for clinical placement as most of them expressed impossibility of maintenance of social distancing in campus, and statement provided by students of government and non-government institute was found statistically significant (1.95±0.98 vs 2.25±1.24, p < 0.0001; 3.94± 1.20 vs 3.65±1.29, p 0.0002). And similar views were expressed in a study conducted among medical students.^[27]^Crowded accomodation facility and lack of cleanliness in hostels of government medical colleges as well as more patient burden in hospital might be the crucial reasons of difference.^[28]^

## Conclusion

COVID-19 pandemic has imposed enormous challenges on medical education worldwide. Undergraduate medical students of both government and non-government institutions of Bangladesh expressed their concern about disruption of medical education, losing of clinical skill, increased level of anxiety and stress, and changing aspect of future career plan. Regarding imposed financial burden, opportunities for meaningful learning, possibility of maintenance of social distancing and fear of getting infected whenever return to clinical placement, significant differences were observed between two groups. Innovative transformations are expected to overcome the impact of COVID-19 pandemic on medical education of Bangladesh.

## Data Availability

All data are available on request

## Acknowledgement (s)

The authors gratefully acknowledge the contributions of 3^rd^, 4^th^and 5^th^ year students of studied medical colleges.

